# Mitochondrial dysfunction drives natural killer cell dysfunction in systemic lupus erythematosus

**DOI:** 10.1101/2025.01.28.25321013

**Authors:** Natalia W. Fluder, Morgane Humbel, Emeline Recazens, Alexis A. Jourdain, Camillo Ribi, George C. Tsokos, Denis Comte

**Affiliations:** Department of Medicine, Division of Immunology and Allergy, Lausanne University Hospital, University of Lausanne, 1011 Lausanne, Switzerland; Department of Medicine, Beth Israel Deaconess Medical Center, Harvard Medical School, Boston, USA; Department of Immunobiology, University of Lausanne, 1066 Epalinges, Switzerland; Department of Medicine, Division of Internal Medicine, Lausanne University Hospital, University of Lausanne, 1011 Lausanne, Switzerland

## Abstract

**Objective:** Systemic lupus erythematosus (SLE) is a chronic autoimmune disease characterized by immune dysregulation and widespread inflammation. Natural killer (NK) cells, essential for immune surveillance, exhibit profound dysfunction in SLE, including impaired cytotoxicity and cytokine production. However, the mechanisms underlying these abnormalities remain poorly understood. This study investigates how the accumulation of dysfunctional mitochondria due to defective mitophagy contributes to NK cell impairment in SLE and explores strategies to restore their function.

**Methods:** Mitochondrial structure and function in NK cells from SLE patients (n=104) and healthy controls (n=104) were assessed using flow cytometry, transmission electron microscopy, and proteomics. Mitophagy-related gene expression was quantified by RT-qPCR. The effects of Urolithin A, a mitophagy activator, and hydroxychloroquine (HCQ) on mitochondrial recycling and NK cell function were evaluated *in vitro*.

**Results:** SLE NK cells exhibited accumulation of enlarged, dysfunctional mitochondria, impaired lysosomal acidification, and increased cytosolic mitochondrial DNA leakage, consistent with defective mitophagy. Proteomic and transcriptional analyses revealed downregulation of key mitophagy-related genes. These abnormalities were associated with diminished NK cell effector functions, including reduced degranulation and cytokine production. *In vitro*, treatment with Urolithin A enhanced mitophagy, improved mitochondrial and lysosomal function, and restored NK cell effector responses. HCQ was also associated with partial recovery of mitochondrial recycling and NK cell function.

**Conclusion:** These findings identify mitochondrial dysfunction and impaired mitophagy as major contributors to NK cell abnormalities in SLE. By uncovering a novel immunometabolic mechanism, this offers new insight into SLE pathogenesis and highlights potential therapeutic strategies targeting mitochondrial quality control.

## INTRODUCTION

Systemic lupus erythematosus (SLE) is a multi-systemic inflammatory autoimmune disease of unknown etiology, which mainly affects women of childbearing age. SLE is characterized by a breakdown in immune tolerance resulting in the production of autoantibodies, immune complexes and autoreactive cells, which subsequently lead to organ damage (1). Although the contribution of different peripheral blood mononuclear cells (CD4+ T, CD8+ T cells and B cells) to the pathogenesis SLE has been widely examined, the precise mechanisms responsible underlying tolerance breakdown remain elusive (2, 3).

Natural Killer (NK) cells represent a group of innate immune cells that play a pivotal role in the interface between the innate and adaptive immune system (4). These cytotoxic lymphocytes are essential for immune surveillance, recognition of healthy cells and defense against viral infections and transformed cells. However, their contribution to autoimmune diseases like SLE remains poorly understood (4). Numerous studies have highlighted NK cell dysfunction in SLE, including reduced numbers in peripheral blood, impaired cytokine production (TNFα, IFNy), reduced cytotoxicity, and defective antibody-dependent cellular cytotoxicity (ADCC) (5–8). These dysregulations may contribute to SLE disease progression by disrupting NK cell interaction with plasma cells, contributing to the persistence of autoantibody-producing cells (9, 10). Additionally, compromised NK cell function may affect therapeutic responses, as monoclonal antibody treatments like rituximab rely on ADCC, a mechanism mediated by functional NK cells (11, 12).

Emerging evidence point to mitochondrial dysfunction as a key driver of immune cell dysregulation in SLE. T cells from SLE patients exhibit abnormal mitochondrial metabolism, including increased mitochondrial mass (megamitochondria), mitochondrial hyperpolarization, and ATP depletion, which contribute to aberrant activation, oxidative stress, and inflammatory signaling—all central features of SLE pathogenesis (13, 14). Moreover, mitophagy, the mitochondrial quality control process, is impaired in SLE T cells, with RAB4A overexpression disrupting lysosomal degradation, leading to the accumulation of damaged mitochondria and persistent immune activation (13–17). Given the dependence of NK cell function on metabolic fitness, we hypothesized that similar mitochondrial abnormalities might contribute to NK cell dysfunction in SLE (13, 18). Additionally, we investigated whether enhancing mitochondrial quality control through pharmacological mitophagy activation could restore mitochondrial integrity and improve NK cell function in SLE.

In the present study, we performed a comprehensive analysis of the mitochondrial homeostasis in NK cells from SLE patients, compared to matched healthy controls (HC). Our aim was to determine how mitochondrial abnormalities affect NK cell function and contribute to immune dysregulation in SLE. Furthermore, we explored the relationship between mitophagy defects and NK cell impairment, providing new insights into the metabolic basis of NK cell dysfunction in SLE and potential therapeutic avenues to restore mitochondrial integrity.

## MATERIALS AND METHODS

### Study design

This observational cross-sectional study aimed to analyze mitochondrial homeostasis in NK cells from patients with SLE compared to matched HC. The objective was to elucidate how immunometabolic abnormalities contribute to NK cell dysfunction and SLE pathogenesis. A total of 104 SLE patients were included based on the 1997 revised American College of Rheumatology (ACR) classification criteria and/or the Systemic Lupus International Collaborating Clinics (SLICC) criteria (28, 29). Patients were enrolled from the Division of Immunology and Allergy at Centre Hospitalier Universitaire Vaudois (CHUV). All participants, including heathy controls, were part of the Swiss Systemic Lupus Erythematosus Cohort Study (SSCS) (30). Written Informed consent was obtained from all individuals, and the study was approved by the Institutional Review Board (SwissEthics 2017-01434), in compliance with the Declaration of Helsinki. Characteristics of the SLE patients included in this study are provided in Table 1.

**Table 1.** Demographic and clinical characteristics of SLE patients included in the study. *Other immunomodulatory drugs include: azathioprine, belimumab, cyclosporine, etanercept, methotrexate, mycophenolate mofetil, rituximab, tacrolimus.

| CHARACTERISTICS |  | N (%) |
| --- | --- | --- |
| **Age, years** |  |  |
|  | Median (SD) | 42 (12.9) |
|  | Range | 20-80 |
| **Sex** |  |  |
|  | Female | 92 (88.5) |
|  | Male | 12 (11.5) |
| **SLE disease activity** |  |  |
|  | Inactive (SLEDAI: 0-3) | 48 (44.5) |
|  | Moderate (SLEDAI: 4-10) | 29 (29.3) |
|  | Active (SLEDAI>10) | 23 (23.2) |
| **Treatment** |  |  |
|  | Naïve | 8 (7.7) |
|  | Prednisone | 57 (54.8) |
|  | HCQ | 74 (71.5) |
|  | HCQ alone | 27 (26) |
|  | Other immunomodulatory drugs * | 44 (42.3) |

#### PBMC isolation & cell culture

Peripheral blood mononuclear cells (PBMCs) were enriched by density gradient centrifugation FICOLL 400 (Merck, Switzerland). The PBMCs were cryopreserved in liquid nitrogen for downstream analyses.

Cells were cultured in RPMI (Gibco; Life Technologies) containing 10% heat inactivated fetal bovine serum (FBS; Institut de Biotechnologies Jacques Boy), 100 IU/ml penicillin and 100 μg/ml streptomycin (Bio Concept), referred to as complete RPMI (cRPMI).

### Flow cytometry

PBMCs from SLE patients and HC were thawed in PRMI containing 20% FBS, centrifuged, resuspended in cRPMI, and pelleted. Cells were stained with Live/Dead-APC-H7 or PB (Invitrogen) and cell surface antibodies: CD3-BUV737or APC-H7, CD56-BUV395, in parallel to different fluorescent probes specific to mitochondrial fitness, superoxides and lysosomal acidification.

#### NK cell mitochondrial fitness

PBMCs were incubated for 20min at 37°C with the cell-permeable fluorescent probes MitoTracker Green and MitotTacker Red (Invitrogen). MitoTracker Green measured mitochondrial mass, while MitoTracker Red assessed the mitochondrial membrane potential (ΔΨ_m_). Cells stained with MitoTracker Red were fixed BD CellFIX™ before analysis on LSR Fortessa™ (BD Bioscience)

#### NK cell mitochondrial superoxides

PBMCs were incubated for 15 min at 37°C, in presence of the MitoSOX fluorescent probe (Invitrogen). PBMCs stained with MitoSOX were analysed on LSR Fortessa™ (BD Bioscience).

#### NK cell lysosomal acidification

PBMCs were incubated for 30-45 min at 37°C, with LysoTracker (Invitrogen) and LysoSensor (Invitrogen) probes to assess lysosomal number and pH, respectively. PBMCs stained with LysoTracker and LysoSensor were analysed on LSR Fortessa™ (BD Bioscience).

#### NK cell cytokine production and degranulation

PBMCs were stimulated with or without cytokines (IL-2 and IL-12, 50ng/ml and 20ng/ml respectively) for 6 hours at 37°C. BD GolgiPlug™, BD GolgiStop™ and CD107a-PE were added 4h before readout. After incubation, cells were stained with Live/Dead-APC-H7 and cell surface antibodies: CD3-BUV737, CD4-PB, CD8-BV605, CD19-FITC, CD56-BUV395. After permeabilization with BD Cytofix/Cytoperm™ kit, cells were stained with IFNγ-AF700, TNFα-APC. before analysis on LSR Fortessa™ (BD Bioscience).

### Hydroxychloroquine (HCQ) and Urolithin A

NK cells or PBMCs from HC and SLE patients were incubated overnight at 37°C, in presence of HCQ sulfate (5mg) or Urolithin A (5mg), purchased from Sigma. PBMCs were treated with 100nM of HCQ sulfate or 1μM of Urolithin A whereas isolated NK cells were stimulated with 1μM of HCQ sulfate or 10μM of Urolithin A.

### NK cell isolation

Natural killer (NK) cells were enriched from SLE and healthy PBMCs using the human NK isolation kit (Miltenyi) with negative labeling. The labeled PBMCs were processed through the AutoMACS® ProSeparator (Miltenyi Biotec).

### NK cell cytosol/mitochondria fragmentation

NK cells were isolated from SLE and HC PBMCs. Cytosolic, mitochondrial, and whole-cell extract fractions were separated using the cytosol/mitochondria fragmentation kit (Abcam) according to the manufacturer’s instructions. Briefly, isolated NK cells were washed in PBS and pelleted by centrifugation at 600g for 5 min. The cell pellet was resuspended in the cytosol extraction buffer and incubated on ice for 10 min, followed by homogenization with a syringe. The homogenate was centrifuged at 1000g for 10 min, and the supernatant, containing the cytosolic fraction, was collected. This supernatant was further centrifuged at 10000g for 30 min. Both supernatant and cell pellet resulting from the last centrifugation were saved as cytosolic fraction and mitochondria, respectively. The mitochondria were resuspended in the mitochondrial extraction buffer and saved as the mitochondrial fraction.

The whole cells extract was obtained by centrifugating NK cells at the initial step, without fractionation. DNA was extracted from all the obtained fractions (cytosolic, mitochondrial, whole cell) using the QIAamp DNA Mini Kit (Qiagen) according to the manufacturer’s instructions.

### Quantification of mitochondrial DNA (mtDNA) content

The mtDNA content in different NK subcellular fractions was quantified using qPCR targeting NADH deshydrogenase 2 (ND2) and D-Loop (displacement loop). Amplifications were performed using PowerTrack SYBR Green and TaqMan Universal PCR Master Mixes (Life Technologies Europe BV). Data were acquired with the QuantStudio™ 6 Flex Real-Time PCR System (ThermoFisher Scientific). Quantification of the mtDNA content in the cytosol of healthy and SLE NK cells was assessed following the basic protocol 2 as previously described (18). Results were normalized to Alu repeat sequence. Primers for qPCR were ordered from Microsynth and are detailed in supplementary materials

### Mitophagy genes expression

RNA was extracted from NK cells of SLE and HC using the RNeasy Plus Mini Kit (Qiagen) according to the manufacturer’s instructions. Complementary DNA was synthesized, and real-time qPCR was performed with SuperScript™ III Platinum™ SYBR™ Green One-Step qRT-PCR Kit (Life Technologies). The expression of mitophagy-related genes including light chain 3B (LC3B), Lysosome-associated membrane protein 2 (LAMP2), PTEN Induced Kinase 1 (PINK1), Parkinson disease 2 (PARK2), class III phosphatidylinositol 3-kinase (PIKC3C), GABA Type A Receptor Associated Protein Like 1 (GABARAPL1), Unc-51-like kinase 1 (ULK1), Beclin 1 (BECN1) genes was assessed using the QuantStudio™ 6 Flex Real-Time PCR System (ThermoFisher Scientific). Results were normalized to the expression b-actin. Primers were ordered from Microsynth (supplementary data).

### Transmission electron microscopy (TEM)

TEM was performed on NK cells isolated from the peripheral blood of SLE (n=5) patients and HC (n=5). Detailed procedures are provided in the online supplemental materials.

### Proteomics analyses

Liquid Chromatography-Mass Spectrometry (LC-MS/MS) analyses were performed on NK cells isolated from the peripheral blood of SLE patients (n=4) and HC (n=4). Detailed procedures are provided in the online supplemental materials. All raw MS data, along with processed raw output tables, are publicly available via the ProteomeXchange data repository (www.proteomexchange.org) with the accession code PXD059825

### Statistics

Statistical analyses were performed using GraphPad Prism (version 10). Details of the statistical tests and sample sizes for each experiment are provided in the corresponding figure legends.

For comparisons between two groups with non-normal distributions and paired data sets the Mann-Whitney U test was used. For comparisons between two groups with non-normal distributions and not paired data sets, the Wilcoxon signed-rank test was performed. Normality was assessed via the Shapiro-Wilk test. For multiple group comparisons with non-normal distributions, the Kruskal-Wallis test was applied, with p-values adjusted using Dunn’s method. For multiple group comparisons with normal distributions, one-way ANOVA was used, with p-values adjusted using the Sidak’s method. For multiple group comparisons with normal distributions and not paired data sets, Mixed effects analysis was used, with p-values adjusted using the Turkey’s method. Two-way ANOVA was used for comparisons involving two independent variables, with p-values adjusted using Sidak’s or Tukey’s methods as appropriate. A p-value <0.05 was considered statistically significant.

## RESULTS

### NK cells from SLE patients accumulate enlarged dysfunctional mitochondria

The numbers of NK cells are reduced in patients with SLE and exhibit impaired degranulation and altered cytokine production. Given that mitochondrial dysfunction has been demonstrated in SLE CD8+ T cells, we sought to investigate whether similar abnormalities contribute to NK cell dysfunction by assessing their mitochondrial fitness (9, 14, 19–21). Mitochondrial function was evaluated using flow cytometry to measure mitochondrial mass, membrane potential, and activity. SLE NK cells exhibited a significant increase in mitochondrial mass (figure 1A) and mitochondrial membrane potential (figure 1B), the latter being reflecting the increase in mitochondrial content. However, normalization of membrane potential to mitochondrial mass revealed a reduction in mitochondrial activity (figure 1C), after 1h of resting. This early time point was chosen to capture initial changes in mitochondrial function. To confirm the persistence of these changes, we reassessed mitochondrial mass in SLE NK cells, after 24h resting period. Although increased mitochondrial mass was observed in other PBMC populations such as B and T cells, this alteration was most pronounced in NK cells from SLE patients compared to HC (supplementary figure 1A). We next investigated whether mitochondrial mass alterations in SLE NK cells correlated with disease activity. Our cohort was divided into three groups: healthy, SLE inactive (SLEDAI ≤ 4), and SLE active (SLEDAI > 4) individuals. Mitochondrial mass (MTK green MFI) and activity (MTK red/green MFI) were assessed across the groups. Mitochondrial mass was significantly increased in relation to disease activity, while mitochondrial activity showed a decreasing trend with increasing severity (figure 1D). To determine whether these functional abnormalities correspond to structural changes, we performed transmission electron microscopy (TEM) analysis on NK cells from SLE patients and HC. TEM revealed that the area occupied by mitochondria was significantly larger in SLE NK cells compared to controls (figure 1E, F). By analyzing 50 TEM images per group, we quantified both mitochondrial area and number. TEM confirmed an increase in mitochondrial size in SLE NK cells, while qPCR analysis showed that the mitochondrial genome remained the same, as indicated by the ratio of mitochondrial to genomic DNA (figure 1G). These findings demonstrate that NK cells from SLE patients accumulate enlarged but dysfunctional mitochondria, suggesting impaired mitochondrial clearance.

**Figure 1:**
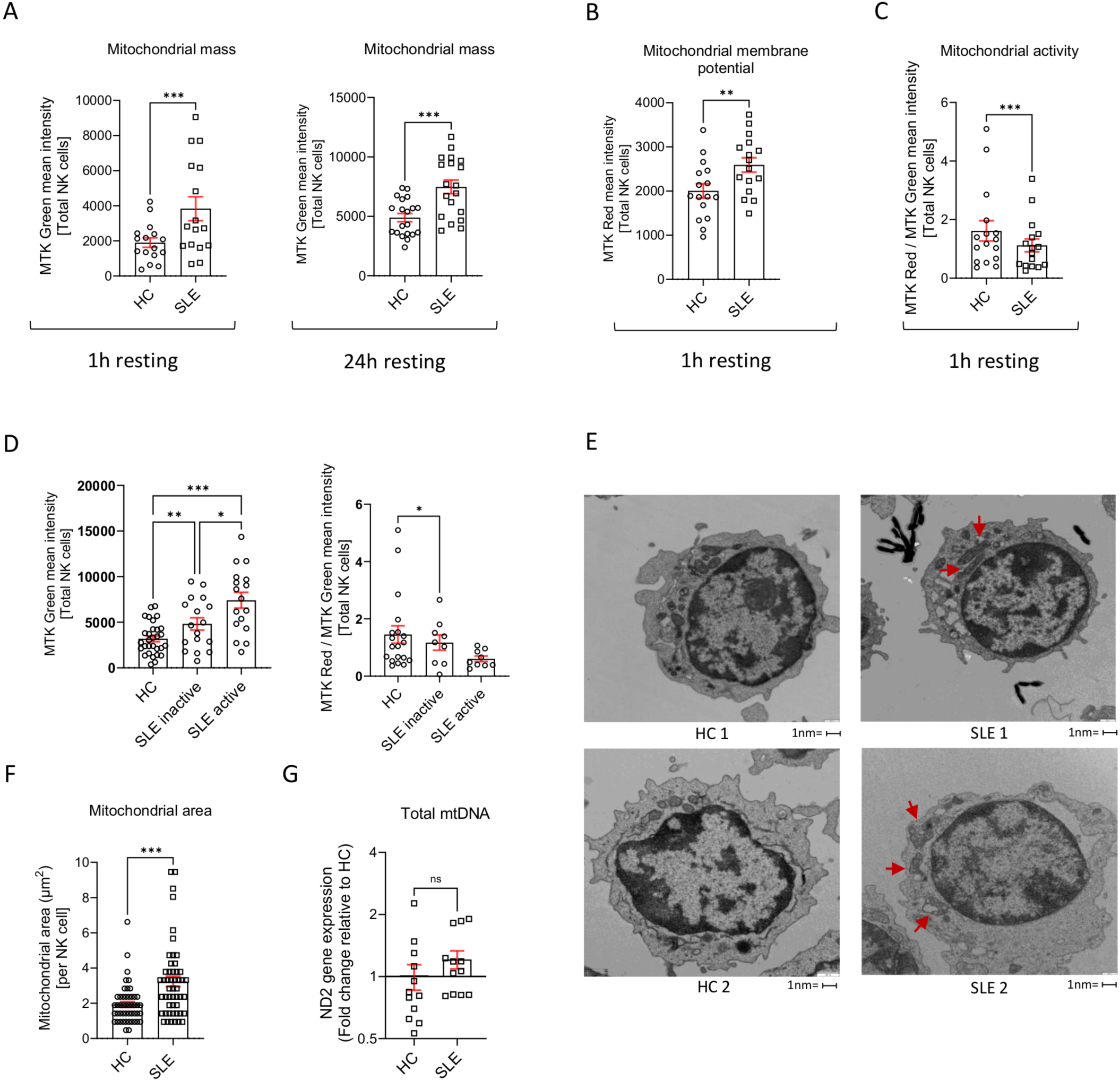
NK cells from SLE patients accumulate enlarged dysfunctional mitochondria. (A) Mitochondrial mass, (B) membrane potential, and (C) activity in NK cells from SLE patients and HC, assessed at baseline by flow cytometry using the MFI of MitoTracker Green, MitoTracker Red, and their ratio, respectively. *p<0.05, by Wilcoxon test. (D) Mitochondrial mass and activity in NK cells from SLE and HC stratified by disease activity score (SLEADAI). *p<0.05, by Mixed-effects analysis. (E) NK cells were isolated from the peripheral blood of HC (n=5) and SLE patients (n=5) and processed for transmission electron microscopy (TEM). Representative TEM images of NK cells from HC and SLE patients at x500 magnification. Red arrows depict examples of the abnormal mitochondria. (F) Mitochondrial area per NK cell was measured on 10 different TEM images per individual from HC (n=5) and SLE patients (n=5) using the grid overlay technique. Each symbol represents a single NK cell. *p<0.05, by Wilcoxon test. (G) NK cells isolated from HC (n=12) and SLE patients (n=12) were analyzed by qPCR on whole cell extracts at baseline. Relative abundance of total mitochondrial DNA (mtDNA) was quantified using the expression of the ND2 gene and normalized to their mean within HC samples. A Log2 fold change (FC) >0.5 or <-0.5 was considered significant, by Wilcoxon test. HC. MFI, mean fluorescence intensity; ND2, NADH dehydrogenase 2 gene.

### Defective lysosomal acidification drives mitochondrial dysfunction in SLE NK cells

To further investigate the mechanisms underlying mitochondrial dysfunction in SLE NK cells, we performed a quantitative proteomic analysis focusing our data analysis on proteins associated with mitochondrial homeostasis. Proteomic profiling revealed increased expression of proteins related to mitochondrial fragmentation (OPA3), suppression of mitophagy (SLC39A7), ER protein retention (KDELR2), negative regulation of NK cell function (NFATC4), mRNA degradation (CNOT10), activation of the NLRP3 inflammasome (APOC3) and metabolites flow through the mitochondrial channel (VDAC1). Conversely, we observed a reduction in proteins key for mitochondrial function (MT-ATP6), energy homeostasis in ER (G6PC3), mitochondrial clearance (MARCHF5) and cellular redox balance, involved in the switching-on of mitophagy (MSRB2) (figure 2A, B). Gene ontology (GO) enrichment analysis also revealed an upregulation of pathways involved in response to DNA damage and DNA metabolic process, along with a downregulation of genes associated with vesicle acidification and trafficking (figure 2C). These findings pointed to potential disruptions in lysosomal function, a critical component of vesicle trafficking. To investigate this, we assessed lysosomal trafficking and acidification in SLE NK cells using flow cytometry. Fluorescent probes specific for lysosomal number and pH revealed that, while the lysosomal number in SLE NK cells was comparable to HC (figure 2D), the lysosomal pH was significantly more alkaline in SLE NK cells (figure 2E). Notably, this alkalinization tended to increase with disease activity in SLE NK cells (figure 2E), suggesting a progressive impairment in lysosomal function. These results suggest that impaired lysosomal acidification may contribute to the functional deficits observed in SLE NK cells, providing new insights into disease pathology. To explore whether lysosomal acidification defects could drive mitochondrial abnormalities in health controls, we treated healthy NK cells with bafilomycin A1, a V-ATPase inhibitor. Bafilomycin A1 treatment resulted in increased mitochondrial mass (figure 2F) and inhibited lysosomal acidification, reproducing the mitochondrial phenotype observed in SLE NK cells (figure 2G). Moreover, bafilomycin A1 (100nM) treatment led to the extrusion of mitochondrial DNA (mtDNA) into the cytosol of healthy NK cells after overnight incubation (figure 2H). These findings establish a critical link between defective lysosomal acidification and mitochondrial dysfunction in NK cells. Impaired lysosomal function likely contributes to the accumulation of enlarged, dysfunctional mitochondria and the cytosolic release of mtDNA, further exacerbating immune dysregulation in SLE.

**Figure 2:**
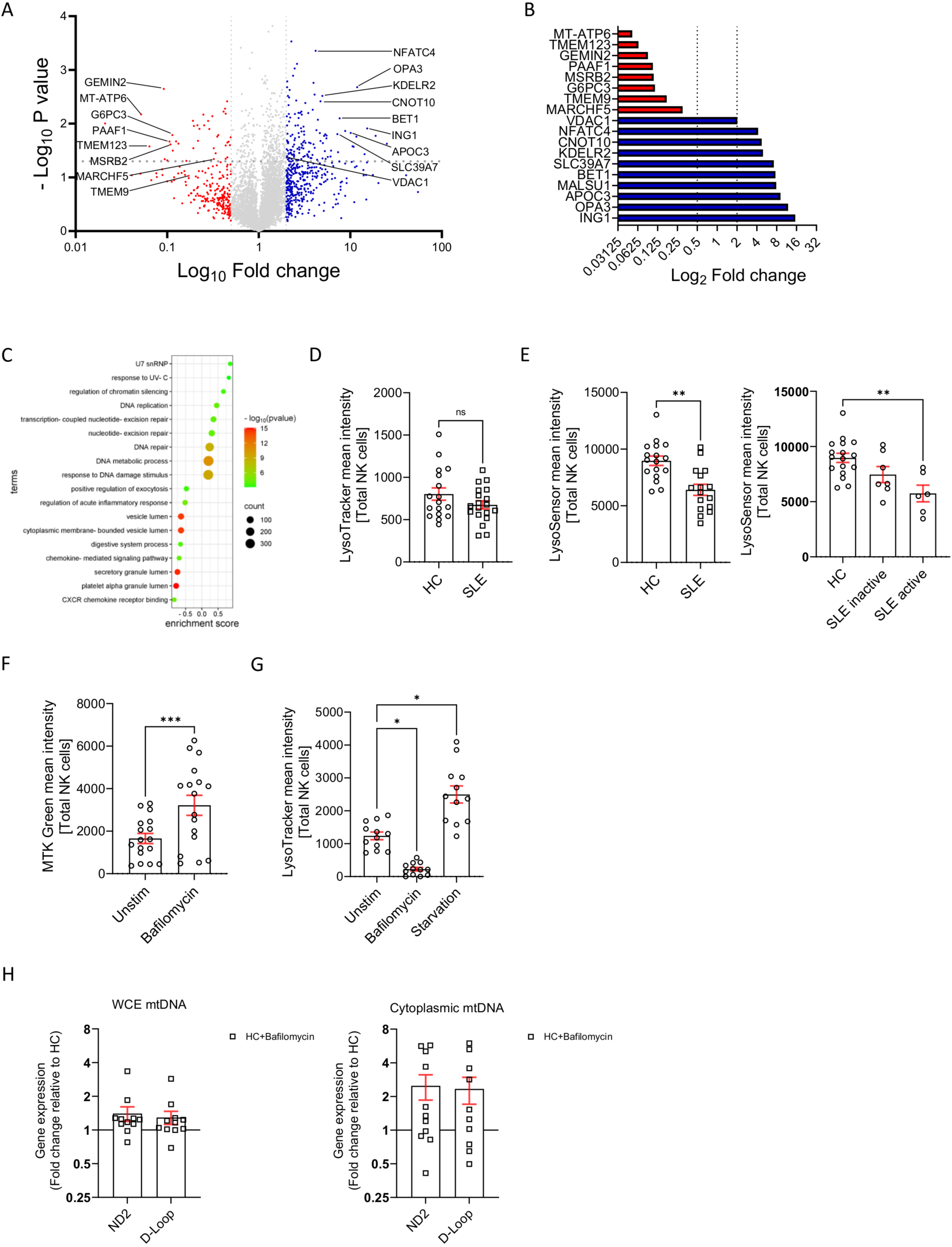
Defective lysosomal acidification drives mitochondrial dysfunction in SLE NK cells. (A) LC-MS–based proteomics was performed on NK cells isolated from the peripheral blood of HC (n=4) and SLE patients (n=4). The volcano plot shows the protein expression levels in NK cells from SLE patients versus HC. Differentially expressed proteins (Log2 fold change [FC] >0.5 or <-0.5) are highlighted in red (upregulated) and blue (downregulated), based on four experiments. (B) Selected significantly upregulated (red) and downregulated (blue) proteins in NK cells from SLE patients compared to HC (p<0.05 and Log2 FC >0.5 or <-0.5), based on four experiments. (C) Gene ontology enrichment analysis of differentially expressed proteins in NK cells from SLE patients compared to HC at baseline. (D, E) Lysosomal number (D) and lysosomal pH (E, left panel) in NK cells from SLE patients and HC were assessed by flow cytometry using the mean fluorescence intensity (MFI) of LysoTracker and LysoSensor probes, respectively. *p<0.05, by Wilcoxon test. (E, right panel) Lysosomal pH in NK cells from SLE and HC was further stratified according to disease activity score (SLEADAI). *p<0.05, by Mixed-effects analysis. (F, G) NK cells from HC were activated overnight in the presence or absence of bafilomycin (100 nM) or subjected to starvation (serum-free RPMI). Mitochondrial mass (F) and lysosomal number (G) in total NK cells were assessed by flow cytometry using the MFI of MitoTracker Green and LysoTracker, respectively. *p<0.05, by Friedman and Wilcoxon test. (H) NK cells from HC (n=11) were activated overnight in the presence or absence of bafilomycin (100 nM), then fractionated by differential ultracentrifugation. qPCR was performed on whole-cell extract (WCE), cytosolic, and mitochondrial fractions. Relative abundance of mtDNA in WCE and cytosolic fraction of HC NK cells was assessed using ND2 and D-Loop genes expression. Data were normalized to unstimulated HC samples. A Log2 FC >0.5 or <-0.5 was considered significant. Abbreviations: LC-MS, liquid chromatography-mass spectrometry; mtDNA, mitochondrial DNA; ND2, D-Loop, mitochondrial DNA displacement loop structure gene.

### NK cells from SLE patients harbor functionally and structurally damaged mitochondria with an altered mitochondrial recycling

Mitochondrial function is essential in maintaining a redox homeostasis where mitochondrial reactive oxygen species (mROS) play a central role in immune regulation. An unbalanced mROS production leads to apoptosis, insufficient cellular debris clearance, oxidative damage, diminished ATP production, and immune dysregulation characterized by skewing pro inflammatory signaling in Th17 cells (20, 22). In our study, mitochondria from SLE NK cells displayed increased levels of superoxide (figure 3A), predisposing them to mitochondrial damage, impaired debris clearance, and increased susceptibility to apoptosis. Notably, mitochondrial superoxide levels correlated with disease activity in SLE NK cells (figure 3A). Electron microscopy showed alterations in the ultrastructure of the mitochondria of SLE NK cells, including mitochondrial cristae disorganization (figure 3B). Given that mitochondrial cristae integrity is crucial for maintaining mtDNA compartmentalization and preventing inflammation, we hypothesized that these structural alterations could facilitate the release of mtDNA into the cytosol. To test this, we performed qPCR analysis to quantify the relative abundance of mtDNA in the cytosol and found significantly higher levels in SLE NK cells compared to HC (figure 3C, D), indicating increased mitochondrial membrane permeability and impaired compartmentalization of mtDNA. In parallel, proteomic analysis of SLE NK cells indicated significant disruptions in mitochondrial homeostasis and clearance mechanism. To further investigate these findings, we measured the expression of key mitophagy-related genes in NK cells from SLE patients and HC using RT-qPCR. Since lysosomal trafficking inhibition is known to induce the accumulation of enlarged mitochondria in healthy NK cells, we examined multiple stages of the autophagy pathway to evaluate the mitophagic flux in SLE NK cells. Our results demonstrated a significant reduction in the expression of genes essential for the PINK1-Parkin–dependent macroautophagy pathway, including, LAMP2, PINK1, PARK2, PIKC3C, GABARAPL2 and BECL1 genes (figure 3E). Together, these findings reveal that SLE NK cells harbor mitochondria with both functional and structural damage, driven in part by impaired mitophagy. These defects likely contribute to the accumulation of dysfunctional mitochondria, mtDNA release, and immune dysregulation, highlighting a critical role for mitochondrial and autophagic pathways in SLE NK cells dysfunction.

**Figure 3:**
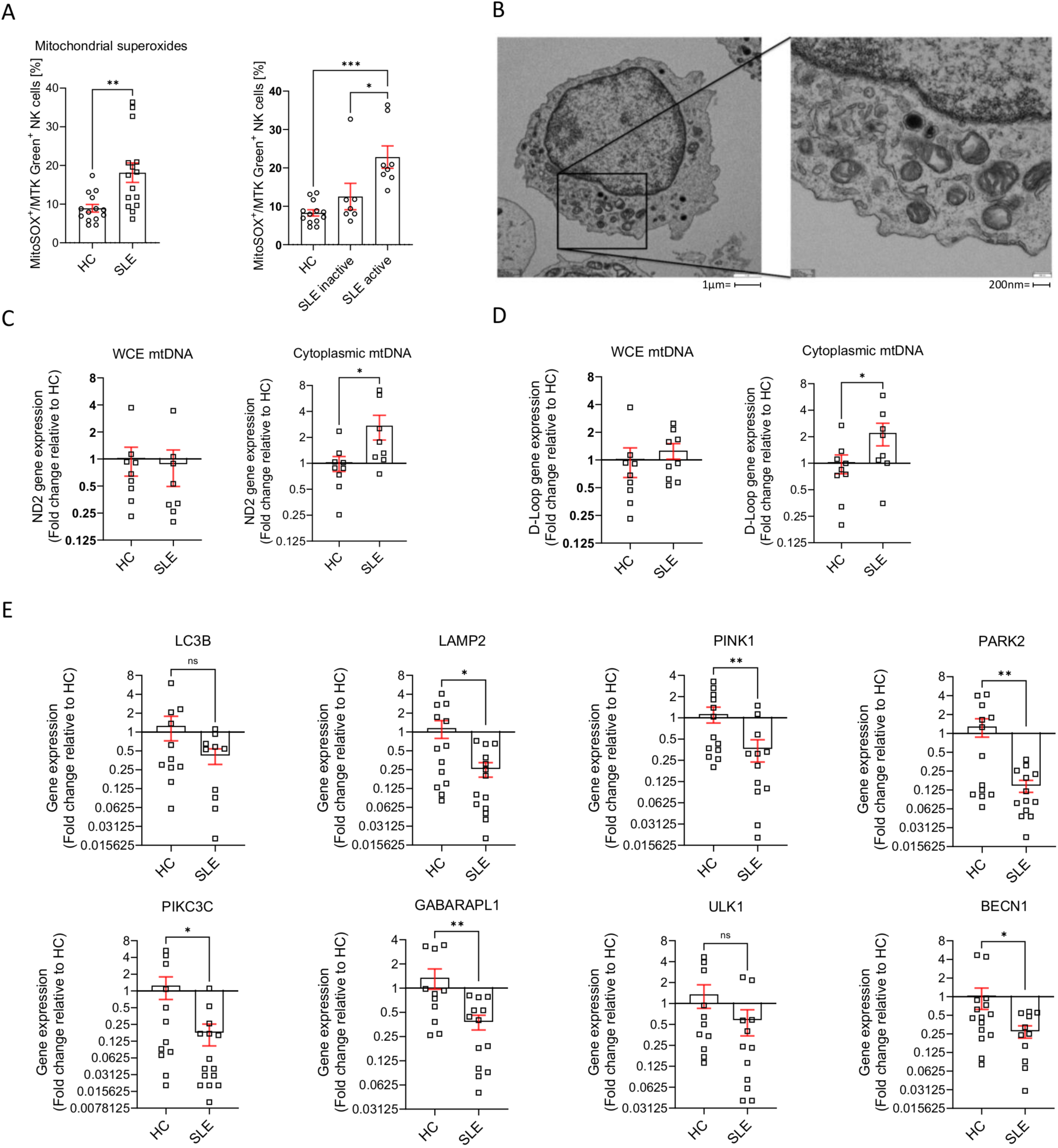
NK cells from SLE patients harbor damaged mitochondria with disrupted ultrastructure and impaired mitophagy. (A) Mitochondrial superoxide levels in NK cells from SLE patients and HC were assessed by flow cytometry, based on the frequency of MitoSOX⁺/MitoTracker Green⁺ cells (left panel). Mitochondrial superoxide levels in NK cells were also stratified according to disease activity score (SLEDAI) (right panel). *p<0.05, by paired t-test (left) and mixed-effects analysis (right). (B) Transmission electron microscopy (TEM) was performed on NK cells from SLE patients. Left panel: representative TEM image at ×5000 magnification showing mitochondrial damage. Right panel: higher magnification image (×15,000) showing mitochondrial ultrastructure abnormalities. (C, D) NK cells were isolated from the peripheral blood of HC (n=10) and SLE patients (n=10), and subcellular fractionation was performed using differential ultracentrifugation. Quantitative PCR (qPCR) was used to assess the relative abundance of mitochondrial DNA (mtDNA) in whole-cell extracts (WCE), cytosolic, and mitochondrial fractions. Expression of ND2 (C) and D-Loop (D) genes was normalized to the mean of HC samples. A Log₂ fold change (FC) >0.5 or <–0.5 was considered significant; *p<0.05, by Mann–Whitney test. (E) Mitophagy-related gene expression (LC3B, LAMP2, PINK1, PIK3C3, GABARAPL2, ULK1, BECN1, and PARK2) in NK cells from HC (n=12) and SLE patients (n=12) was assessed at baseline by RT-qPCR. Expression values were normalized to the mean expression in HC. A Log₂ FC >0.5 or <–0.5 was considered significant; *p<0.05, by Mann–Whitney test. Abbreviations: MitoSOX, mitochondrial superoxides; MTK; LC3B, light chain 3B; LAMP2, lysosome-associated membrane protein 2; PINK1, PTEN Induced Kinase 1; PARK2, Parkinson disease 2; PIKC3C, class III phosphatidylinositol 3-kinase; GABARAPL1, GABA type A Receptor Associated Protein Like 1; ULK1, Unc-51-like kinase 1; BECN1, Beclin 1.

### Pharmacological activation of mitophagy by Urolithin A restores mitochondrial integrity, lysosomal acidification and NK cell function in SLE

We and others have previously demonstrated that NK cell function is severely impaired in SLE, characterized by defective production of IFNγ and TNFα and impaired degranulation, as indicated by decreased CD107a expression (6, 9, 23). To test whether restoring mitochondrial homeostasis could rescue NK cell function, we used Urolithin A, a natural mitophagy activator known to enhance mitochondrial clearance and improve cellular bioenergetics. This approach allowed us to assess whether improving endogenous mitochondrial turnover could functionally reverse NK cell dysfunction in SLE. SLE NK cells were activated overnight *in vitro* with IL-2 + IL-12. The addition of Urolithin A to the culture enhanced degranulation (CD107a) (figure 4A), increased cytokine production (IFNγ, TNFα) (figure 4B, C), partially corrected mitochondrial mass (figure 4D), and restored lysosomal acidification (figure 4E, F), compared to cells not exposed to Urolithin A. These findings support the hypothesis that mitochondrial dysfunction is a key driver of NK cell impairment in SLE and that targeting mitophagy can restore NK cell function.

**Figure 4:**
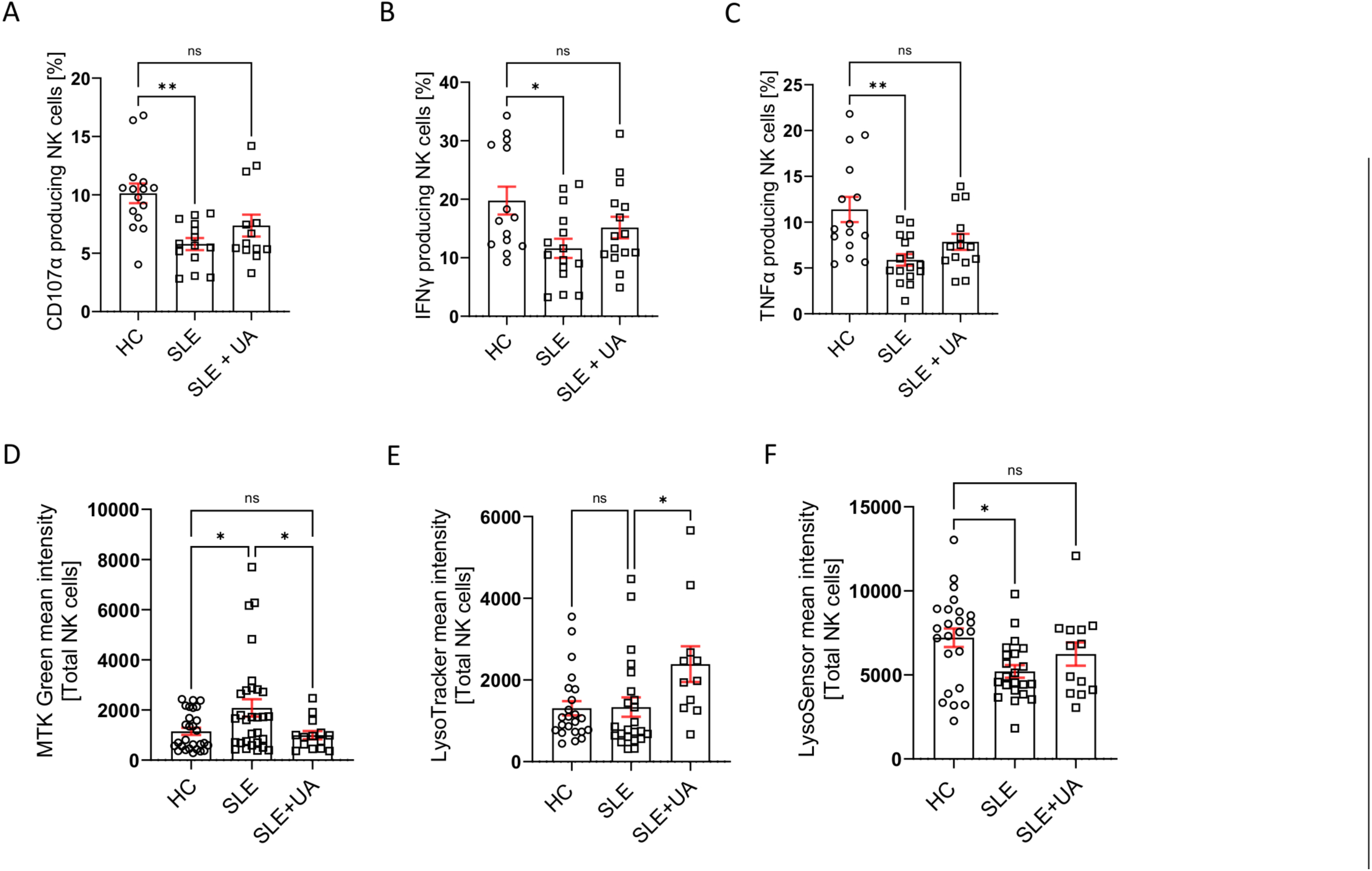
Pharmacological activation of mitophagy by Urolithin A (UA) restores mitochondrial integrity, lysosomal acidification and NK cell function in SLE. NK cells from SLE patients and HC were activated overnight with IL-2 and IL-12 in the presence or absence of UA (UA, 1 uM). (A) Degranulation and (B, C) cytokine production in NK cells were assessed by flow cytometry, based on the frequency of CD107α+ (A), IFNγ+ (B), and TNFα+ (C) cells. *p<0.05, **p<0.01, ***p<0.001, by Kruskal-Wallis test. (D) Mitochondrial mass in total NK cells was assessed by flow cytometry using the MFI of MitoTracker Green. *p<0.05, by Ordinary one-way ANOVA. (E, F) Lysosomal number (E) and lysosomal pH (F) were assessed by flow cytometry at baseline using the MFI of fluorescent probes (LysoTracker for lysosomal number; LysoSensor for lysosomal pH). *p<0.05, **p<0.01, ***p<0.001, by Kruskal-Wallis test.

### Hydroxychloroquine restores NK cell function in SLE patients

SLE NK cells exhibit mitochondrial dysfunction, impaired mitophagy, and diminished effector functions. To explore whether HCQ, a widely used therapeutic in SLE, could restore these cellular processes, we evaluated its effects on NK cell activation, mitochondrial recycling, and mitophagy-related gene expression (supplementary figure 2). *In vitro* treatment of activated (IL-2+IL-12) SLE NK cells with HCQ reversed their reduced cytotoxicity and cytokine production, restoring these functions to levels observed in HC (figure 5A, B, C). To investigate whether HCQ could also address mitochondrial dysfunction, we examined its effects on lysosomal acidification. Our data demonstrated that HCQ effectively acidifies lysosomes in SLE NK cells (figure 5D, E). Moreover, we investigated the impact of HCQ on mtDNA release into the cytosol, a hallmark of mitochondrial dysfunction in SLE NK cells. Stimulation with HCQ significantly reduced the relative levels of cytosolic mtDNA in NK cells from HC, compared to untreated cells (figure 5F, G).

**Figure 5:**
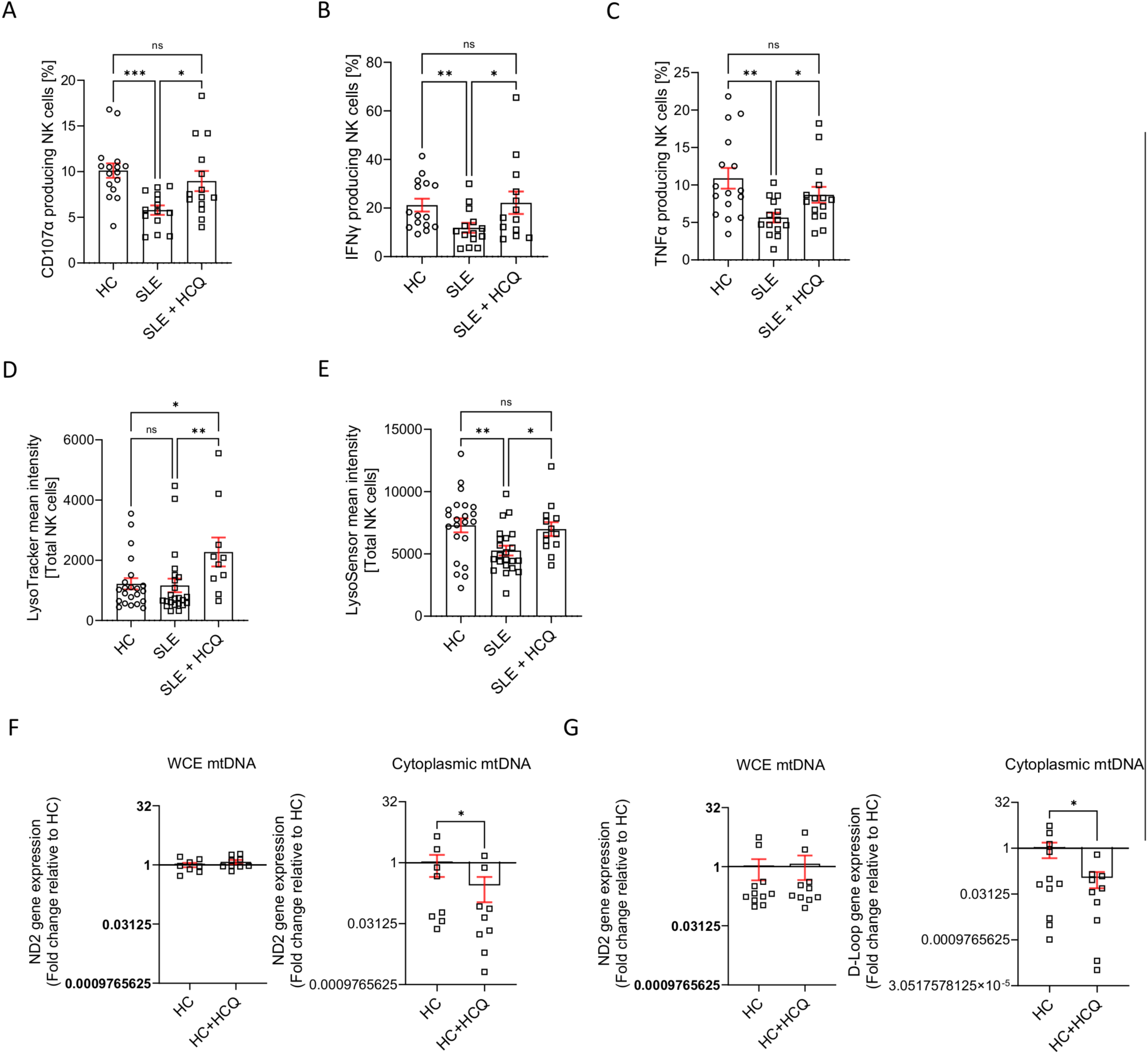
Hydroxychloroquine (HCQ) restores the cell function in NK cells from SLE patients. NK cells from HC and SLE patients were activated overnight with IL-2 and IL-12 in the presence or absence of HCQ (100 nM). (A) Degranulation and (B, C) cytokine production (IFNγ, TNFα) in NK cells from HC and SLE patients were assessed by flow cytometry based on the frequency of CD107α+ (A), IFNγ+ (B), and TNFα+ (C) cells. *p<0.05, **p<0.01, ***p<0.001, by Kruskal-Wallis test. (D, E) Lysosomal number (D) and lysosomal pH (E, left panel) in NK cells from SLE patients and HC were assessed by flow cytometry using the mean fluorescence intensity (MFI) of LysoTracker and LysoSensor probes, respectively. *p<0.05, **p<0.01, ***p<0.001, by Kruskal-Wallis test. (F, G) NK from HC (n=10) were activatedovernight with or without HCQ (1 µM) and then fractionated by differential ultracentrifugation. qPCR was performed on whole-cell extracts, cytosolic, and mitochondrial fractions. The relative abundance of cytosolic mtDNA was assessed using the expression of the ND2 (F) and D-Loop (G) genes. A Log2 fold change (FC) >0.5 or <-0.5 was considered significant, by Wilcoxon test.

### Hydroxychloroquine restores the mitochondrial recycling in NK cells from SLE patients

Finally, we examined the effect of HCQ on the expression of key mitophagy-related genes. Overnight incubation with HCQ restored the expression of LC3B, LAMP2, PINK1, PARK2, PIK3C3, GABARAPL2, ULK1 and BECL1 in SLE NK cells to levels comparable to those of HC (figure 6). Together, these findings demonstrate that HCQ not only restores NK cell effector functions but also addresses mitochondrial and autophagic dysregulation in SLE, highlighting its potential therapeutic benefits in managing NK cell dysfunction.

**Figure 6:**
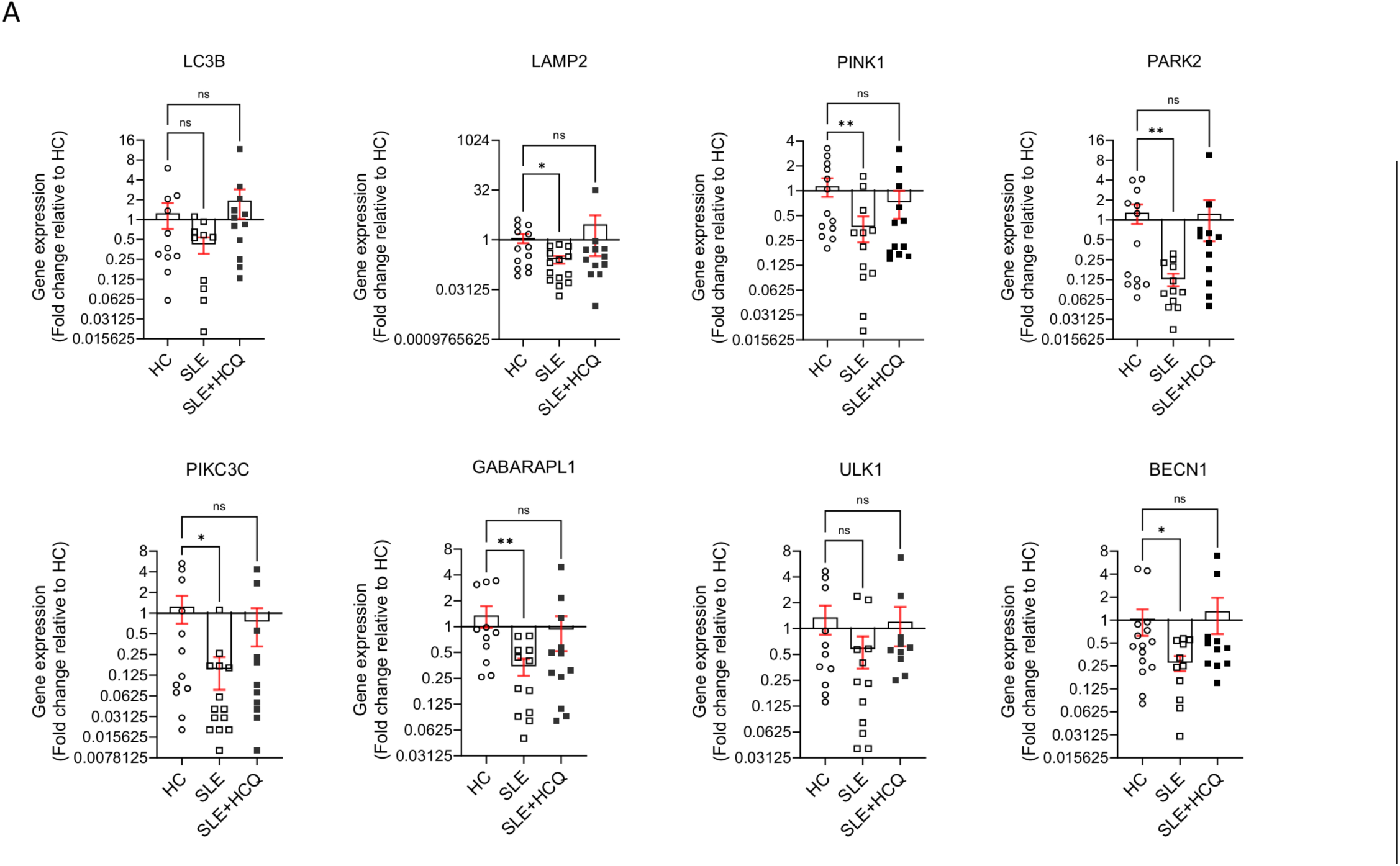
Hydroxychloroquine (HCQ) restores the mitochondrial recycling in NK cells from SLE patients. (A) NK cells from SLE patients (n=11) and HC were activatedovernight with or without HCQ (1 µM), followed by RT-qPCR analysis. The relative expression of mitophagy-related genes (LC3B, LAMP2, PINK1, PARK2, PIKC3C, GABARAPL2, ULK1, and BECL1) was assessed and normalized to HC samples. A Log2 FC >0.5 or <-0.5 was considered significant, by Kruskal-Wallis test. Abbreviations: LC3B, light chain 3B; LAMP2, lysosome-associated membrane protein 2; PINK1, PTEN Induced Kinase 1; PARK2, Parkinson disease 2; PIKC3C, class III phosphatidylinositol 3-kinase; GABARAPL1, GABA type A Receptor Associated Protein Like 1; ULK1, Unc-51-like kinase 1; BECN1, Beclin 1

## DISCUSSION

NK cells are essential components of the immune system, playing key roles in antiviral defense, tumor surveillance, and immune regulation through cytotoxic activity and cytokine production (5). In SLE, NK cells exhibit profound dysfunction, including reduced cytotoxicity, impaired degranulation, and diminished cytokine production (e.g., IFNγ and TNFα) (6–8). These defects have significant implications for SLE pathogenesis. First, the inability to eliminate virally infected and transformed cells increases susceptibility to infections and malignancies in SLE patients (24). Second, the failure to clear autoreactive cells, such as antibody-producing plasma cells, CD4+ T cells, and dendritic cells, exacerbates autoantibody production and contributes to inflammation, immune complex deposition, and tissue damage (25–27). Third, NK cell dysfunction compromises antibody-dependent cellular cytotoxicity (ADCC), undermining the efficacy of monoclonal antibody therapies such as rituximab, particularly in severe cases like lupus nephritis (11, 12).

To elucidate the mechanisms underlying NK cell dysfunction in SLE, we focused on their immunometabolic regulation, with particular attention to mitochondrial fitness and recycling. Our findings revealed significant mitochondrial abnormalities in SLE NK cells, including increased mitochondrial mass, hyperpolarized membranes, and reduced activity, indicative of severe mitochondrial dysfunction. Additionally, defects in mitophagy, a critical mitochondrial quality control pathway, were observed, with SLE NK cells exhibiting reduced expression of genes involved in PINK1-Parkin– dependent macroautophagy pathway (LAMP2, PINK1, PARK2, PIKC3C, GABARAPL2, BECL1) and impaired lysosomal acidification.

Alterations in these pathways suggest a profound impairment in mitochondrial quality control. Reduced expression of PINK1 and PARK2 prevent efficient identification of dysfunctional mitochondria, allowing their prolonged accumulation within cells. The downregulation ofGABARAPL1, PIK3C3 and BECN1 disrupts autophagosome formation and cargo selection, impairing the initiation of mitophagy. Furthermore, insufficient LAMP2 expression may lead to defective autophagosomes-lysosome fusion, ultimately impairing lysosomal degradation of damaged mitochondria. These abnormalities result in the accumulation of dysfunctional mitochondria leading to oxidative stress and the extrusion of mitochondrial DNA (mtDNA) into the cytosol, a key driver of inflammation through activation of cGAS and type I interferon responses, contributing to the amplification of autoimmune mechanisms. Together, these findings suggest that mitochondrial dysfunction and impaired mitophagy are central contributors to NK cell abnormalities and broader immune dysregulation in SLE. The ability of Urolithin A to restore mitochondrial integrity and NK cell function provides indicate that enhancing mitochondrial quality control is sufficient to rescue NK cell effector functions. This supports the hypothesis that defective mitochondrial recycling, rather than intrinsic NK cell signaling defects, is a primary driver of their dysfunction in SLE.

HCQ is widely used in SLE to reduce disease activity, prevent flares, and decrease corticosteroid dependency, with additional benefits such as thrombotic risk reduction and improved pregnancy outcomes (21, 22). At the molecular level, HCQ accumulates in lysosomes, modulating pH and affecting key processes like autophagy, antigen presentation, and cytokine production. While high doses (5–50 µM) can impair lysosomal function and induce toxicity, our study demonstrated that low, clinically relevant, doses (0.1–1 µM) effectively restore NK cell function without cytotoxic effects. Given HCQ’s multifaceted mechanisms of action, it is unclear whether its observed effects on mitochondrial function are direct or secondary to its modulation of lysosomal activity and immune signaling. Additionally, a significant proportion of patients in our cohort were already receiving HCQ treatment, raising the possibility that any *in vitro* effects may not fully replicate *in vivo* responses. While our results suggest that HCQ partially restores mitochondrial homeostasis and reduces mtDNA release, further studies will be needed to determine whether these effects are due to direct modulation of mitophagy or secondary to its immunomodulatory properties.

Several limitations should be acknowledged. First, the cross-sectional nature of this study limits our ability to determine whether mitochondrial dysfunction is a primary driver of NK cell impairment in SLE or a secondary consequence of chronic immune activation. Future studies should include longitudinal analyses to better define the temporal relationship between mitochondrial abnormalities and NK cell function. Second, while our findings suggest an impairment in mitophagy, our study primarily relied on gene expression analyses to assess autophagy-related pathways. Given that autophagy is a dynamic process, further studies assessing protein markers such as LC3B and p62/SQSTM1, as well as functional mitophagy flux assays, will be needed to confirm these findings. Third, we recognize that the MTK Red/Green ratio provides a relative measure of mitochondrial membrane potential and mass but does not directly assess mitochondrial ATP production. While Seahorse assays would provide additional insights into mitochondrial bioenergetics, their feasibility in NK cells isolated from frozen PBMCs is limited. Fourth, the use of Bafilomycin A1 as a tool to block lysosomal acidification requires careful interpretation. Bafilomycin A1 can indirectly affect mitochondrial dynamics by preventing mitochondrial degradation, which may complicate the assessment of mitophagy. We now explicitly acknowledge this limitation in our revised discussion. Finally, we recognize that frozen PBMCs may exhibit metabolic alterations that could influence mitochondrial readouts. However, given that both SLE and HC NK cells were processed under identical conditions, we believe that the observed differences are biologically meaningful. Nevertheless, future studies using freshly isolated NK cells will be required to fully validate these findings.

In conclusion, our study identifies mitochondrial dysfunction and impaired mitophagy as central drivers of NK cell abnormalities in SLE. These defects contribute to oxidative stress, mtDNA release, and chronic immune activation, providing new insights into how NK cell metabolism influences disease pathogenesis. While HCQ appears to partially restore mitochondrial homeostasis, its precise mechanism of action in this context remains unclear. Future research should explore targeted approaches to modulate mitochondrial quality control mechanisms in NK cells, which may provide novel therapeutic strategies to restore immune balance and improve clinical outcomes in SLE.

## Data Availability

All data produced in the present study are available upon reasonable request to the authors

## Data availability

The raw data supporting the conclusions of this article will be made available by the authors, without undue reservation.

## Author Contributions

NF, DC: study design and data analysis. NF, MH, ER: conducting experiments, data acquisition and analysis. NF, CR, DC: recruitment of SLE patients and HC. NF, DC: writing and editing of manuscript. MH, GTC, ER, AAJ, CR: critical reading of the manuscript. All authors contributed to the article and approved the submitted version.

## Funding

This work was funded by a grant from the Swiss National Science Foundation Ambizione PZ00P3_173950 (DC), a grant from the Novartis Foundation 173950 (DC), a grant from the National Science Foundation 310030_200796 (AAJ) and PHS NIH R01AI148161 (GCT).

## Conflict of Interest

The authors declare that the research was conducted in the absence of any commercial or financial relationships that could be construed as a potential conflict of interest.

## Acknowledgments

We thank Professor Giuseppe Pantaleo for his support. The personnel of the vaccine and immunotherapy center for the isolation of SLE PBMC cells, and study nurse Emmanuelle Paccou along with the physicians of the division of immunology and allergy, for their help with patient recruitment. We are also grateful to the Swiss Systemic Lupus Erythematosus Cohort Study (SSCS) for their support. Additionally, we acknowledge the staff of the UNIL electron microscopy and proteomic facilities for performing NK cell imaging and protein expression analysis.

## Notes

### Competing Interest Statement

The authors have declared no competing interest.

### Author Declarations

Ethics committee/IRB of Lausanne University Hospital (Commission cantonale d'ethique de la recherche sur l'etre humain - CER-VD, Lausanne, Switzerland) gave ethical approval for this work

### Summary of Updates

This version of the manuscript has been substantially revised to improve clarity, precision, and alignment with reviewer feedback. The title and abstract have been updated to better reflect the central finding that mitochondrial dysfunction drives natural killer (NK) cell impairment in systemic lupus erythematosus (SLE). The introduction and discussion were revised to more clearly position mitophagy defects as a core mechanism of NK cell dysregulation, while tempering claims about hydroxychloroquine (HCQ) to reflect its association with, rather than proof of, functional restoration. A new section has been added describing the effects of pharmacological mitophagy activation using Urolithin A as a surrogate for mitochondrial rescue. Figures and legends have been edited for consistency, and gene and protein names have been standardized throughout the text. Minor typographical and stylistic corrections were also made

## References

1. Tsokos GC. Systemic lupus erythematosus. N Engl J Med. 2011;365(22):2110–21.

2. Nashi E, Wang Y, Diamond B. The role of B cells in lupus pathogenesis. Int J Biochem Cell Biol. 2010;42(4):543–50.

3. Suárez-Fueyo A, Bradley SJ, Tsokos GC. T cells in Systemic Lupus Erythematosus. Curr Opin Immunol. 2016;43:32–8.

4. Moretta A, Marcenaro E, Parolini S, Ferlazzo G, Moretta L. NK cells at the interface between innate and adaptive immunity. Cell Death Differ. 2008;15(2):226–33.

5. Spada R, Rojas JM, Barber DF. Recent findings on the role of natural killer cells in the pathogenesis of systemic lupus erythematosus. J Leukoc Biol. 2015;98(4):479–87.

6. Park YW, Kee SJ, Cho YN, Lee EH, Lee HY, Kim EM, et al. Impaired differentiation and cytotoxicity of natural killer cells in systemic lupus erythematosus. Arthritis Rheum. 2009;60(6):1753–63.

7. Tsokos GC, Rook AH, Djeu JY, Balow JE. Natural killer cells and interferon responses in patients with systemic lupus erythematosus. Clin Exp Immunol. 1982;50(2):239–45.

8. Rook AH, Tsokos GC, Quinnan GV, Jr., Balow JE, Ramsey KM, Stocks N, et al. Cytotoxic antibodies to natural killer cells in systemic lupus erythematosus. Clin Immunol Immunopathol. 1982;24(2):179–85.

9. Humbel M, Bellanger F, Fluder N, Horisberger A, Suffiotti M, Fenwick C, et al. Restoration of NK Cell Cytotoxic Function With Elotuzumab and Daratumumab Promotes Elimination of Circulating Plasma Cells in Patients With SLE. Front Immunol. 2021;12:645478.

10. Takeda K, Dennert G. The development of autoimmunity in C57BL/6 lpr mice correlates with the disappearance of natural killer type 1-positive cells: evidence for their suppressive action on bone marrow stem cell proliferation, B cell immunoglobulin secretion, and autoimmune symptoms. J Exp Med. 1993;177(1):155–64.

11. Merrill JT, Neuwelt CM, Wallace DJ, Shanahan JC, Latinis KM, Oates JC, et al. Efficacy and safety of rituximab in moderately-to-severely active systemic lupus erythematosus: the randomized, double-blind, phase II/III systemic lupus erythematosus evaluation of rituximab trial. Arthritis Rheum. 2010;62(1):222–33.

12. Rovin BH, Furie R, Latinis K, Looney RJ, Fervenza FC, Sanchez-Guerrero J, et al. Efficacy and safety of rituximab in patients with active proliferative lupus nephritis: the Lupus Nephritis Assessment with Rituximab study. Arthritis Rheum. 2012;64(4):1215–26.

13. Caza TN, Fernandez DR, Talaber G, Oaks Z, Haas M, Madaio MP, et al. HRES-1/Rab4-mediated depletion of Drp1 impairs mitochondrial homeostasis and represents a target for treatment in SLE. Ann Rheum Dis. 2014;73(10):1888–97.

14. Chen PM, Katsuyama E, Satyam A, Li H, Rubio J, Jung S, et al. CD38 reduces mitochondrial fitness and cytotoxic T cell response against viral infection in lupus patients by suppressing mitophagy. Sci Adv. 2022;8(24):eabo4271.

15. Fernandez DR, Telarico T, Bonilla E, Li Q, Banerjee S, Middleton FA, et al. Activation of mammalian target of rapamycin controls the loss of TCRzeta in lupus T cells through HRES-1/Rab4-regulated lysosomal degradation. J Immunol. 2009;182(4):2063–73.

16. Caza TN, Talaber G, Perl A. Metabolic regulation of organelle homeostasis in lupus T cells. Clinical Immunology. 2012;144(3):200–13.

17. Zhao L, Hu X, Xiao F, Zhang X, Zhao L, Wang M. Mitochondrial impairment and repair in the pathogenesis of systemic lupus erythematosus. Front Immunol. 2022;13:929520.

18. Bryant JD, Lei Y, VanPortfliet JJ, Winters AD, West AP. Assessing Mitochondrial DNA Release into the Cytosol and Subsequent Activation of Innate Immune-related Pathways in Mammalian Cells. Curr Protoc. 2022;2(2):e372.

19. Quintero-González DC, Muñoz-Urbano M, Vásquez G. Mitochondria as a key player in systemic lupus erythematosus. Autoimmunity. 2022;55(8):497–505.

20. Lu Z, Tian Y, Bai Z, Liu J, Zhang Y, Qi J, et al. Increased oxidative stress contributes to impaired peripheral CD56(dim)CD57(+) NK cells from patients with systemic lupus erythematosus. Arthritis Res Ther. 2022;24(1):48.

21. Morel L. Immunometabolism in systemic lupus erythematosus. Nature Reviews Rheumatology. 2017;13(5):280–90.

22. Yang J, Yang X, Zou H, Li M. Oxidative Stress and Treg and Th17 Dysfunction in Systemic Lupus Erythematosus. Oxid Med Cell Longev. 2016;2016:2526174.

23. Schleinitz N, Vély F, Harlé J-R, Vivier E. Natural killer cells in human autoimmune diseases. Immunology. 2010;131(4):451–8.

24. Mace EM, Orange JS. Emerging insights into human health and NK cell biology from the study of NK cell deficiencies. Immunol Rev. 2019;287(1):202–25.

25. Schuster IS, Wikstrom ME, Brizard G, Coudert JD, Estcourt MJ, Manzur M, et al. TRAIL+ NK cells control CD4+ T cell responses during chronic viral infection to limit autoimmunity. Immunity. 2014;41(4):646–56.

26. Cruz-González DJ, Gómez-Martin D, Layseca-Espinosa E, Baranda L, Abud-Mendoza C, Alcocer-Varela J, et al. Analysis of the regulatory function of natural killer cells from patients with systemic lupus erythematosus. Clin Exp Immunol. 2018;191(3):288–300.

27. Yang Y, Day J, Souza-Fonseca Guimaraes F, Wicks IP, Louis C. Natural killer cells in inflammatory autoimmune diseases. Clin Transl Immunology. 2021;10(2):e1250.

28. Tan EM, Cohen AS, Fries JF, Masi AT, McShane DJ, Rothfield NF, et al. The 1982 revised criteria for the classification of systemic lupus erythematosus. Arthritis Rheum. 1982;25(11):1271–7.

29. Petri M, Orbai AM, Alarcón GS, Gordon C, Merrill JT, Fortin PR, et al. Derivation and validation of the Systemic Lupus International Collaborating Clinics classification criteria for systemic lupus erythematosus. Arthritis Rheum. 2012;64(8):2677–86.

30. Ribi C, Trendelenburg M, Gayet-Ageron A, Cohen C, Dayer E, Eisenberger U, et al. The Swiss Systemic lupus erythematosus Cohort Study (SSCS) – cross-sectional analysis of clinical characteristics and treatments across different medical disciplines in Switzerland. Swiss Medical Weekly. 2014;144(3132):w13990.

