## Supplemental material for "Mitochondrial dysfunction drives natural killer cell dysfunction in systemic lupus erythematosus"

**SUPPLEMENTARY MATERIALS**

**Transmission electron microscopy (TEM)**Cells were centrifugated at 2500rpm, supernatant washed out and then the cells were fixed in 2.5% glutaraldehyde solution (EMS, Hatfield, PA) and in osmium tetroxide 1% (EMS) with 1.5% of potassium ferrocyanide (Sigma, St. Louis, MO) in phosphate buffer (PB 0.1 M [pH 7.4]) for 1h at RT. The samples were then centrifugated at 5000rpm, supernatant washed out and replaced by distilled water twice. Then the cells were again centrifugated at 5000rpm and the supernatant removed. After that, the cells were dehydrated in acetone solution (Sigma, St Louis, MO, US) at graded concentrations (30%-15 min; 70% - 15 min; 100% - 4x15). This was followed by infiltration in Epon resin (EMS, Hatfield, PA, US) at graded concentrations (Epon 33% in acetone-2h; Epon 66% in acetone-4h; Epon 100%-2x8h) and finally polymerized for 48h at 60°C in an oven. Ultrathin sections of 50 nm were cut using a Leica UC7 (Leica Mikrosysteme GmbH, Vienna, Austria), picked up on a copper slot grid 2x1mm (EMS, Hatfield, PA, US) coated with a polystyrene film (Sigma, St Louis, MO, US). Sections were post-stained with uranyl acetate (Sigma, St Louis, MO, US) 4% in H2O for 10 min, rinsed several times with H2O followed by Reynolds lead citrate in H2O (Sigma, St Louis, MO, US) for 10 min and rinsed several times with H2O. Micrographs were taken with a transmission electron microscope FEI CM100 (FEI, Eindhoven, The Netherlands) at an acceleration voltage of 80kV with a TVIPS TemCamF416 digital camera (TVIPS GmbH, Gauting, Germany).

**Proteomic analyses*Sample preparation and protein digestion*** Washed cell pellets (3-5 10^5^ cells) were lysed in 25 μl miST lysis buffer (1% Sodium deoxycholate, 100mM Tris pH 8.6, 10 mM DTT) and heated for 10min at 75°C. Proteins were digested following a modified version of the iST method (Kulak, et al., 2014) (named miST method). Based on tryptophane fluorescence quantification (Wisniewski, et al., 2015), approximately 30 ug of proteins at 1μg/μl were transferred to new tubes, diluted 1:1 (v:v) with water containing 4mM MgCl_2_ and benzonase (Merck #70746, 100x dil of stock = 250 Units/μl), and incubated for 15 minutes at RT to digest nucleic acids. Reduced disulfides were alkylated by adding ¼ vol. of 160 mM chloroacetamide (32 mM final) and incubating for 45min at RT in the dark. Samples were adjusted to 3 mM EDTA and digested with 0.5 μg Trypsin/LysC mix (Promega #V5073) for 1h at 37°C, followed by a second 1h digestion with an additional 0.5μg of proteases. To remove sodium deoxycholate, two sample volumes of isopropanol containing 1% TFA were added to the digests, and the samples were desalted on a strong cation exchange (SCX) plate (Oasis MCX; Waters Corp., Milford, MA) by centrifugation. After washing with isopropanol 1%TFA, peptides were eluted in 200μl of 80% MeCN, 19% water, 1% (v/v) ammonia, and dried by centrifugal evaporation.

***Liquid Chromatography-Mass Spectrometry analyses*** LC-MS/MS analyses were carried out on a TIMS-TOF Pro (Bruker, Bremen, Germany) mass spectrometer interfaced through a nanospray ion source (“captive spray”) to an Ultimate 3000 RSLCnano HPLC system (Dionex). Peptides were separated on a reversed-phase custom packed 45 cm C18 column (75 μm ID, 100Å, Reprosil Pur 1.9 um particles, Dr. Maisch, Germany) at a flow rate of 250 nl/min with a 2-27% acetonitrile gradient in 93 min followed by a ramp to 45% in 15 min and to 90% in 5 min (total method time: 140 min, all solvents contained 0.1% formic acid). Approximately 0.25 μg of digest were injected.

The data-independent acquisition (DIA) used mostly the instrument parameters reported previously (Meier, et al. 2020). Per cycle, the mass range 400-1200 m/z was covered by a total of 32 windows, each 25 Th wide and a 1/k0 range of 0.3. Collision energy was ramped linearly based uniquely on the 1/k0 values from 20 (at 1/k0=0.6) to 59 eV (at 1/k0=1.6). Two windows were acquired per TIMS scan (100ms) so that the total cycle time was 1.7 s.

***Data processing*** Raw Bruker MS DIA data were processed directly with Spectronaut 16.1 (Biognosys, Schlieren, Switzerland) with the Pulsar engine using the “deep” setting and searching the human SWISSPROT database ([www.uniprot.org](http://www.uniprot.org)) of January 7^th^, 2022 (20’375 sequences). For identification, peptides of 7-52 AA length were considered, cleaved with trypsin/P specificity and a maximum of 2 missed cleavages. Carbamidomethylation of cysteine (fixed), methionine oxidation and N-terminal protein acetylation (variable) were the modifications applied. FDR’s for peptide and protein group identifications were all at 1%. Ion mobility for peptides was predicted using a deep neural network and used in scoring. The library created contained overall 91’819 precursors (71’900 peptides).

Peptide-centric analysis of DIA data was done with Spectronaut 16.1 using the library generated by Pulsar from DIA data. Single hits proteins (defined as matched by one stripped sequence only) were kept in the Spectronaut analysis. Peptide quantitation was based on XIC area, for which a minimum of 1 and a maximum of 3 (the 3 best) precursors were considered for each peptide, from which the median value was selected. Quantities for protein groups were derived from inter-run peptide ratios based on MaxLFQ algorithm (Cox et al 2014). Global normalization of runs/samples was done based on the median of peptides. Overall 6’360 protein groups were identified at 1% FDR.

***Data analysis*** All subsequent analyses were done with the Perseus software package (version 1.6.15.0) (Tyanova et al 2016). Quantities were log2-transformed and contaminants removed. After assignment to conditions, only proteins quantified in at least 2 samples of one condition were kept (6’256 protein groups). After missing values imputation (based on normal distribution using Perseus default parameters), t-tests were carried out among all conditions, with permutation-based FDR correction for multiple testing (Q-value threshold <0.05). Imputed values were later removed. The difference of means obtained from the tests were used for 1D enrichment analysis on associated GO/KEGG annotations as described (Cox and Mann, 2012). The enrichment analysis was also FDR-filtered (Benjamini-Hochberg, Q-val<0.02).

Supplementary figure 1: Specificity of mitochondrial mass increase within PBMCs from SLE patients and healthy controls. Mitochondrial mass (A-D) in NK (A), B- (B), CD8+ T- (C), CD4+T- (D) cells from HC (n=19) versus SLE patients (n=19), assessed at baseline by flow cytometry, using the MFI of MitoTracker Green. All readouts were performed at baseline, after overnight resting. Each symbol represents one individual. *p<0.05, by Wilcoxon test.

Supplementary figure 2: Impact of hydroxychloroquine concentration of NK cells phenotype alteration in healthy controls. Dot plots representing gating on NK cells (CD3-, CD56+) performed during the optimization of our assay measuring the lysosomal number, assessed by flow cytometry, using the MFI LysoTracker. PBMCs from healthy controls were stimulated overnight with ± bafilomycin (100nM), different concentrations of Hydroxychloroquine (HCQ): 0.01, 0.1, 1, 10, 100, 1000 μM or starved.
