## Supplemental Figure 1 for "Mitochondrial dysfunction drives natural killer cell dysfunction in systemic lupus erythematosus"

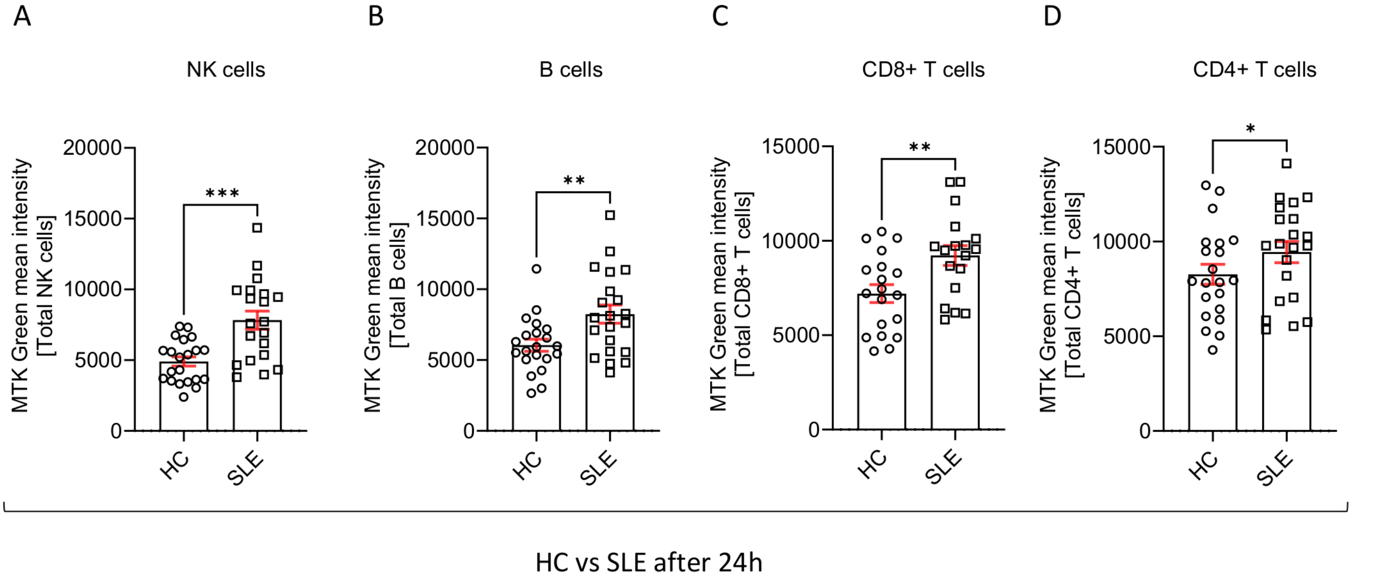


Supplemental figure 1: Specificity of mitochondrial mass increase within PBMCs from SLE patients and healthy controls.
