## Supplemental Figure 2 for "Mitochondrial dysfunction drives natural killer cell dysfunction in systemic lupus erythematosus"

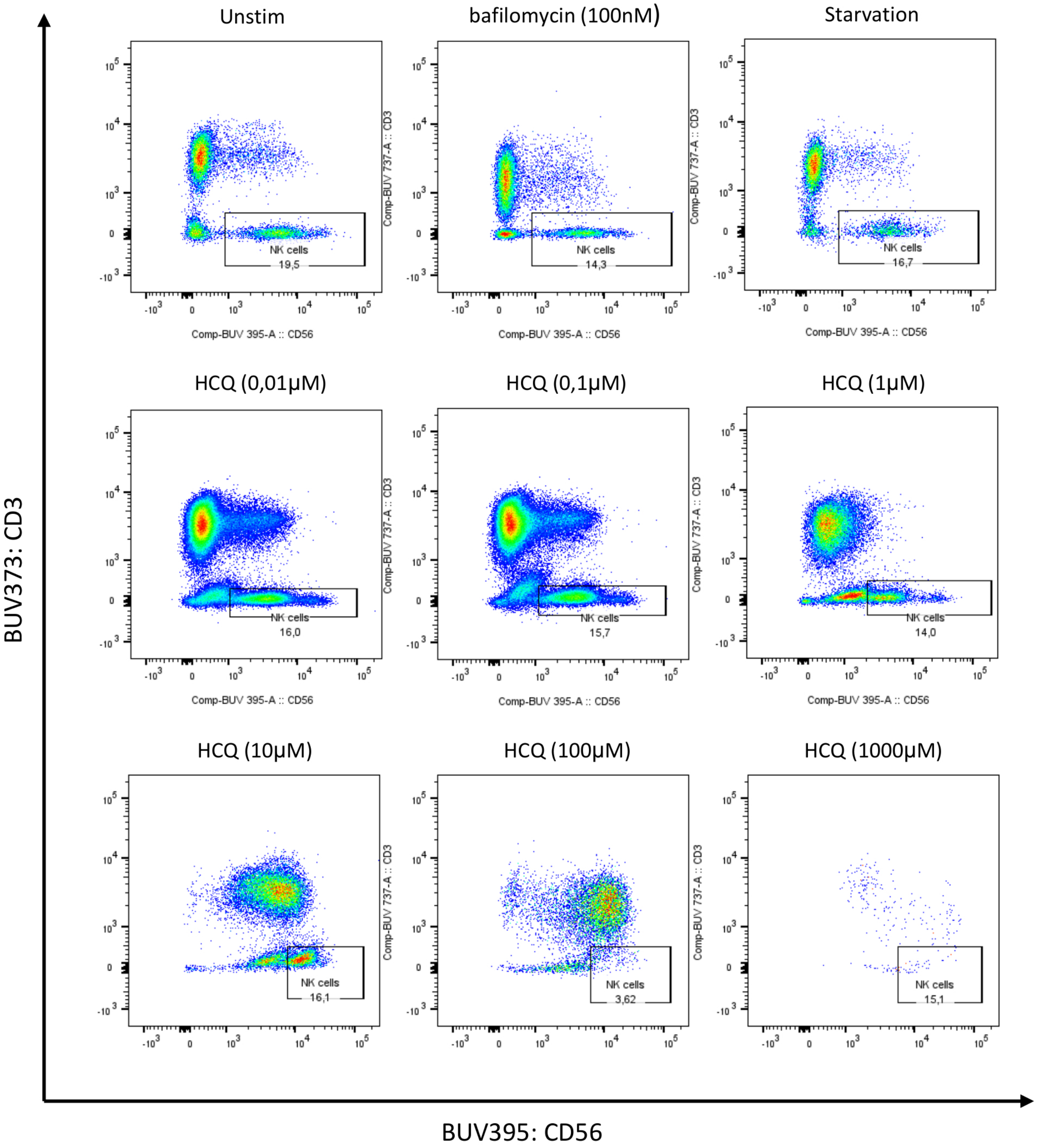


<u>Supplemental figure 2:Impact of hydroxychloroquine concentration of NK cells phenotype alteration in healthy controls.</u>
